# Proportion, patterns and Factors Associated with Diabetic Macular Edema among Diabetes Type 2 Patients attending KCMC Hospital 2023-2024

**DOI:** 10.1101/2025.06.27.25330425

**Authors:** Mutima Nadege, William Makupa, Maria Kisanga

## Abstract

**Objective:** To determine the prevalence and risk factors associated with DME and clinically significant macular edema (CSME) among type 2 diabetes mellitus (T2DM) patients attending Kilimanjaro Christian Medical Centre (KCMC) from August 2023 to June 2024.

**Methods:** A hospital-based cross-sectional study was conducted among 384 T2DM patients at KCMC’s medical and ophthalmology outpatient departments. Data collected included sociol demographic characteristics, clinical history, ocular examinations (including OCT imaging), and biochemical parameters. CSME was defined according to Early Treatment Diabetic Retinopathy Study (ETDRS) criteria. Statistical analyses were performed using STATA version 17. Chi-square tests determined prevalence, while modified Poisson regression identified associated risk factors (p<0.05).

**Results:** The mean age was 63.4±11.1 years; 55.2% were male, and 63.3% were aged over 60. The prevalence of DME was 33.9%, with diffuse macular edema being the most common subtype (40.8%). Significant risk factors for DME included hypertension, elevated HbA1c, reduced glomerular filtration rate (GFR), abnormal BMI, and longer DM duration. For CSME, elevated total cholesterol, hypertension, and BMI were independently associated.

**Conclusion:** DME affects one in three T2DM patients at KCMC and is strongly associated with modifiable cardio metabolic risk factors. Strengthening early detection and comprehensive diabetes management—including control of blood pressure, glucose, lipids, and body weight—is essential to reduce vision-threatening complications.

## Introduction

Diabetic macular edema (DME) is a vision-threatening complication of diabetes mellitus characterized by fluid accumulation in the macula, which can lead to significant visual impairment and blindness if left untreated. Diabetes currently affects an estimated 537 million adults globally—a figure projected to rise to 643 million by 2030. Approximately one-third of these individuals develop diabetic retinopathy, and around 5–11% of patients with type 2 diabetes mellitus (T2DM) go on to experience DME, especially when evaluated using optical coherence tomography (OCT(1,2).The burden is particularly high in low- and middle-income countries (LMICs), where inadequate healthcare infrastructure and limited screening programs exacerbate disease progression and outcomes.

In Sub-Saharan Africa (SSA), DME affects between 5% and 14% of individuals with diabetes, with diabetic retinopathy prevalence ranging from 7% to 62% (3).In Tanzania, adult diabetes prevalence is approximately 9%, and locally reported maculopathy—including DME—affects between 5% and 17% of diabetic patients at presentation (4).Despite this, phenotypic patterns of DME and associated risk factors remain poorly characterized in this region.

OCT-based morphological studies typically identify three common patterns of DME: diffuse retinal thickening being the leading (present in about 43–62% of affected eyes), cystoid macular edema (37–60%), and serous retinal detachment (7–21%), with the latter often seen in more severe disease associated with high HbA1c and advanced retinopathy (5– 7)Modifiable risk factors consistently associated with DME and clinically significant macular edema (CSME) across global and regional studies include poor glycemic control (elevated HbA1c), hypertension, longer diabetes duration, dyslipidemia, obesity, renal dysfunction (proteinuria or reduced eGFR), and albuminuria (8–10)Regional data further underscore the link between total cholesterol, renal markers. Despite compelling global evidence, region-specific data—particularly from East Africa—is limited. The aim of the study was to evaluate the proportion, Patterns and Factors associated with Diabetic macula edema among type 2 diabetes patients attending KCMC Hospital from August 2023 to June 2024

## Material and methods

This was cross-sectional study conducted at Kilimanjaro Christian Medical Centre (KCMC), Moshi, Tanzania, from August 2023 to June 2024. The study included adult patients (≥□18□years) with a confirmed diagnosis of type□2 diabetes mellitus (T2DM) and diabetic retinopathy (DR), recruited consecutively from the ophthalmology and medical outpatient departments. The study protocol was approved by the Kilimanjaro Christian medical Research Ethics Committee (reference PG121/2023).All participants provided written informed consent before enrollment. Procedures adhered to the Declaration of Helsinki.

Data were collected using structured forms capturing demographics, medical history, examination findings, and laboratory results.

Intraocular pressure measurement was done by Goldman applanation tonometry while Fundoscopic examinations using 90D Volk lenses after pupillary dilation .Macular edema was defined clinically by retinal thickening or hard exudates near the fovea; OCT imaging (Carl Zeiss Primus 200) quantified central subfield macular thickness (CSMT) and macular cube volume, using a threshold of ≥250□µm and cube volume ≥10□mm^3^.

We analyzed the data using Stata version 17 (StataCorp, USA), Chi-square tests assessed proportional differences, while modified Poisson regression identified factors associated with diabetic macula edema.

## RESULTS

A total of 384 participants were included in this study. The mean age of the study participants was 63.4(±11.14) years with 243 (63.3%) of the participants aged above than 60 years. More than half of the study participants, 212 (55.2%) were male and 151 (39.3%) of the participants were overweight. There were 220 (57.3%) participants who had diabetes 5 years and above, while 350(91.1%) of the study participants were on oral anti diabetes treatment. There were 136(32.8%) participants who had hypertension. Of the study participants, 126(32.8%) were alcohol takers, where by 23(6%) were cigarette smokers” **Table”**

**Table 1:** Participants’ background characteristics (N=384)

| Variable | Frequency | Percentage |
| --- | --- | --- |
| **Participants' age in years** |  |  |
| ≤60 | 141 | 36.7 |
| >60 | 243 | 63.3 |
| *Mean(SD)* | *63.4(±11.14)* |  |
| **Sex** |  |  |
| Male | 212 | 55.2 |
| Female | 172 | 44.8 |
| **BMI** |  |  |
| Underweight | 9 | 2.3 |
| Normal | 115 | 29.9 |
| Overweight | 151 | 39.3 |
| Obese | 109 | 28.4 |
| **Duration of diabetes (In years)** |  |  |
| <5 | 164 | 42.7 |
| ≥5 | 220 | 57.3 |
| **Treatment option of diabetes** |  |  |
| Oral anti diabetes | 350 | 91.1 |
| Insulin | 20 | 5.2 |
| Diet | 14 | 3.6 |
| **Hypertension** |  |  |
| No | 248 | 64.6 |
| Yes | 136 | 35.4 |
| **Alcohol intake** |  |  |
| No | 258 | 67.2 |
| Yes | 126 | 32.8 |
| **Cigarette smoking** |  |  |
| No | 361 | 94 |
| Yes | 23 | 6 |

### Laboratory results of the study participants

**“Table 2”,** summarizes laboratory results of the study participants. The mean (SD) of HB1ac was 7.4(±2.79) with 241(62.8%) had high HB1ac, while the median (IQR) of serum creatinine was 80.3(60-98) with 300(78.1%) having normal creatine. Moreover, the median (IQR) of Glomerular filtration rate and Uric acid was 82.3(56.5-96.2) and 248(201-350) respectively, with 270(70.3%) participants having normal GFR and 220(57.3%) were on safe level of uric acid. Additionally, Serum urea had a median (IQR) of 4.7(3.4-6.4) with 300(78.1) having normal serum urea while the Mean (SD) of total cholesterol was 4.7(±1.42) with 230 (59.9%) having normal cholesterols

**Table 2:** Laboratory results of the study participants (N=384)

| Variable | Frequency | Percentage |
| --- | --- | --- |
| **HB1ac** |  |  |
| Normal HB1ac | 143 | 37.2 |
| High HB1ac | 241 | 62.8 |
| *Mean(SD)* | 7.4(±2.79) | |
| **Serum creatinine** |  |  |
| Normal creatinine | 300 | 78.1 |
| Raised creatinine | 84 | 21.9 |
| *Median(IQR)* | 80.3(60-98) |  |
| **Glomerular filtration rate** |  |  |
| Kidney disease | 114 | 29.7 |
| Normal GFR | 270 | 70.3 |
| *Median(IQR)* | 82.3(56.5-96.2) |  |
| **Uric acid** |  |  |
| Safe | 220 | 57.3 |
| Good | 72 | 18.8 |
| Warning level | 24 | 6.3 |
| Danger | 68 | 17.7 |
| *Median(IQR)* | 248(201-350) |  |
| **Serum urea** |  |  |
| Normal Urea | 300 | 78.1 |
| High Urea | 84 | 21.9 |
| *Median(IQR)* | 4.7(3.4-6.4) |  |
| **Total cholesterol** |  |  |
| Normal cholesterol | 230 | 59.9 |
| Border line | 72 | 18.8 |

**Table 3:** Factors associated with diabetic macula edema by Participants Characteristics.

| **Variable** | **CPR(95%CI)** | **P-value** | **APR(95%CI)** | **P-value** |
| --- | --- | --- | --- | --- |
| **Age of participants** |  |  |  |  |
| ≤60 | Ref |  | Ref |  |
| >60 | 0.74(0.56-0.98) | 0.036 | 0.85(0.66-1.11) | 0.242 |
| **Sex** |  |  |  |  |
| Male | Ref |  |  |  |
| Female | 1.02(0.77-1.36) | 0.867 |  |  |
| **Hypertension** |  |  |  |  |
| No | Ref |  | Ref |  |
| Yes | 2.57(1.94-3.39) | <0.001 | 1.43(1.08-1.88) | 0.012 |
| **Duration of diabetes(In years)** |  |  |  |  |
| <5 | Ref |  | Ref |  |
| ≥5 | 2.84(1.96-4.13) | <0.001 | 1.47(1.06-2.03) | 0.021 |
| **BMI** |  |  |  |  |
| Normal | Ref |  | Ref |  |
| Abnormal | 1.52(1.08-2.15) | 0.016 | 1.46(1.11-1.91) | 0.007 |
| **HB1ac** |  |  |  |  |
| Normal HB1ac | Ref |  | Ref |  |
| High HB1ac | 5.83(3.34-10.19) | <0.001 | 3.84(2.27-6.49) | <0.001 |
| **Serum creatinine** |  |  |  |  |
| Normal creatinine | Ref |  | Ref |  |
| Raised creatinine | 2.97(2.32-3.80) | <0.001 | 0.86(0.56-1.31) | 0.484 |
| **Glomerular filtration rate** |  |  |  |  |
| Normal GFR | Ref |  | Ref |  |
| Kidney disease | 3.13(2.39-4.10) | <0.001 | 2.18(1.37-3.49) | 0.001 |
| **Uric acid** |  |  |  |  |
| Normal uric acid level | Ref |  | Ref |  |
| High uric acid level | 2.38(1.84-3.09) | <0.001 | 0.94(0.68-1.30) | 0.715 |
| **Serum urea** |  |  |  |  |
| Normal Urea | Ref |  | Ref |  |
| High Urea | 2.62(2.04-3.36) | <0.001 | 1.36(0.99-1.86) | 0.057 |
| **Total cholesterol** |  |  |  |  |
| Normal cholesterol | Ref |  |  |  |
| High cholesterol | 0.85(0.63-1.14) | 0.264 |  |  |

### Proportion of macula edema by Participants characteristics

The proportion of diabetic macula edema was 33.9% (130/384). In this study, 40.8% (33/130) of the study participants had diffuse macula edema with least of the participants having serous macula edema 2.3% (3/130). “**Figure 3”**

**Figure 1:**
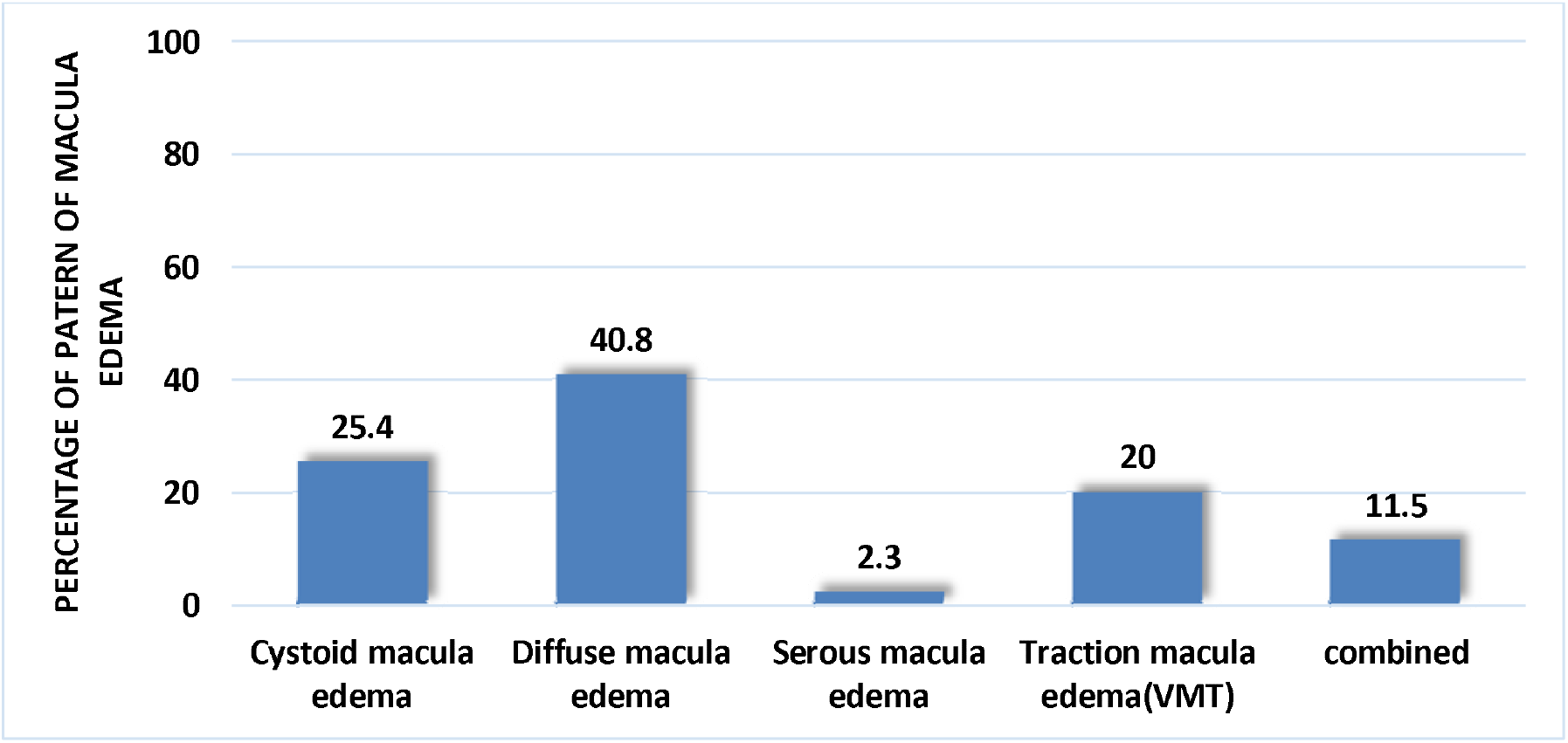
Pattern of macula edema among the study participants (N=130)

### Factors associated with diabetic macula edema by Participants Characteristics

**“Table 4**”, summarizes factors associated with diabetic macula edema, whereby in crude analysis, age of the participant, hypertension, DM duration, BMI, HB1ac, serum creatinine, glomerular filtration rate, uric acid and serum urea, were factors that significantly associated with macula edema.

Participants who aged greater than 60 years were 26% significant less prevalent to have macula edema compared to participants who aged 60 years and below (CPR=0.74; 95%CI (0.56-0.98): P-value=0.0036), while compared to participants without hypertension, participants with hypertension were 2.6 times significant more prevalent of having diabetic macula edema (CPR=2.57 95%CI (1.94-3,39): P-value<0.001). Participants with DM duration 5 years and above were 2.8 time significantly more prevalent of having diabetic macula edema (CPR=2.84; 95%CI (1.96-4.13): P-value<0.001). Compared to participants with normal BMI, participants with abnormal BMI were 52% significantly more prevalent of having diabetic macula edema below (CPR=1.52; 95%CI (1.08-2.14): P-value=0.016), whereby individuals with high HB1ac were 5.8 times significantly more prevalent of having diabetic macula edema compared to individuals normal HB1ac (CPR=5.83; 95%CI (3.34-10.18): P-value<0.001). Participants with raised creatinine were 2.9 times significantly more prevalent of having macula edema compared to individuals with normal creatine (CPR=2.97; 95%CI (2.32-3.80): P-value<0.001). Moreover, individuals with kidney diseases were 3.1 times significantly more prevalent of having diabetic macula edema compared to individuals without kidney disease (CPR=3.13 95%CI (2.39-4.10): P-value<0.001). Compared to individuals with normal uric acid, individuals with high uric acid were 2.4 times significantly more prevalent of having diabetic macula edema (CPR=2.38; 95%CI (1.84-3.09): P-value<0.001) and individuals with high urea were 2.6 time significantly more prevalent of having diabetic macula edema compared to individuals with normal urea (CPR=2.62; 95%CI (2.04-3.36): P-value<0.001).

In adjusted analysis hypertension, DM duration, BMI, HB1ac and glomerular filtration rate were factors that significantly associated with diabetic macula edema. After adjusting for age of the participant, DM duration, BMI, HB1ac, serum creatinine, glomerular filtration rate, uric acid and serum urea, compared to participants without hypertension, participants with hypertension were 42% significant more prevalent of having diabetic macula edema (APR=1.43 95%CI (1.08-1.88): P-value=0.012) while compared to participants with DM duration 5 years and above were 47% significantly more prevalent of having diabetic macula edema (APR=1.47; 95%CI (1.05-2.03): P-value=0.021) after adjusting for age of the participant, hypertension, BMI, HB1ac, serum creatinine, glomerular filtration rate, uric acid and serum urea. Compared to participants with normal BMI, participants with abnormal BMI were 46% significantly more prevalent of having diabetic macula edema (APR=1.46; 95%CI (1.11-1.91): P-value=0.007) after adjusting for age of the participant, hypertension, DM duration, HB1ac, serum creatinine, glomerular filtration rate, uric acid and serum urea. Additionally, after adjusting for age of the participant, hypertension, DM duration, BMI, serum creatinine, glomerular filtration rate, uric acid and serum urea individuals with high HB1ac were 3.8 times significantly more prevalent of having diabetic macula edema compared to individuals normal HB1ac (APR=3.83; 95%CI (2.27-6.49): P-value<0.001) while individuals with kidney diseases were 2.2 times significantly more prevalent of having diabetic macula edema compared to individuals without kidney disease (APR=2.18 95%CI (1.37-3.48): P-value=0.001) after adjusting for age of the participant, hypertension, DM duration, BMI, HB1ac, serum creatinine, uric acid and serum urea.

## Discussion

This study found high proportion of DME (33.9%), a referral nature of our hospital setting might have introduced a selection bias, as patients referred to KCMC are likely to have more severe or advanced stages of diabetes and its complications. This could result in a higher observed prevalence of DME compared to community-based or primary care settings. The majority had poor glycemic control, and a notable minority displayed abnormal kidney function and cholesterol levels. Findings were similar to the study done on Risk Factors and Incidence of Macular Edema after Cataract Surgery described that diabetic Retinopathy and Projection of Burden through found prevalence of poor glycemic control is a major risk factor for the development and progression of diabetic retinopathy and DME(11).

Our study revealed that 33.9% had DME, with diffuse macular edema being the most prevalent type (40.8%). Our findings show that most of DME pattern was diffuse macula edema 40.8%, followed by cystoids macula edema 25.4%, Serous macula edema 2.3%, traction macula edema 20.0% and 11.5% have combined macular edema. The dominant of diffuse macula edema in this study can be explained by the patient at KCMC presented at advanced stage of diabetic retinopathy caused by delayed diagnosis and treatment initiation. The findings vary in a study done in Pakistan found that Spongy 13% have macular Edema, 51% have Cystic Macular Edema, and 76 have Focal macular Edema(12). The different can be explained by different in study design and geographical area.

Also, our findings were similar to the study done in India on the Prevalence and Pattern of Macular Edema in Diabetes found that diffuse retinal thickening in 9 eyes 42.4%, cystoids macular edema in 7 eyes 36.8%, and serous retinal detachment in 15.8% (7). Also, the similarity can be explained by same design and geographical area.

The study identified that kidney disease significantly increased the likelihood of DME by more than twice. Elevated serum creatinine levels and impaired glomerular filtration rate were significantly associated with higher prevalence rates of DME in our study, increasing by 2 times, respectively. Diabetic kidney disease (DKD) and impaired renal function are closely intertwined with diabetic retinopathy and DME due to shared pathophysiological mechanisms involving microvascular damage and systemic metabolic dysfunction. Comprehensive management of DKD and renal function is essential in diabetic patients to mitigate the risk of DME and other microvascular complications The study findings were similar to, kidney diseases as associated with DME in a Study done in Turkey on the prevalence and systemic risk factors of diabetic macular edema. However, the factors were not similar such as alcohol consumption, neuropathy, previous cataract surgery, severity of diabetic retinopathy, and insulin usage as factors associated with DME(6). Despite these differences, the consistent association of kidney disease with DME suggests a strong link between the two across various studies.

Our research revealed that individuals with high HbA1c levels were three times more likely to develop diabetic macular edema. Elevated HbA1c reflects poor long-term glycemic control, a key driver of micro vascular complications in diabetes. Tight glycemic control through pharmacological interventions and lifestyle modifications is crucial in preventing and delaying the onset and progression of DME among diabetic patients.

These findings are consistent with a study conducted in the USA on the prevalence and risk factors for diabetic macular edema, which also found a significant association between elevated levels of hemoglobin and DME, (13). This similarity suggests that poor glycemic control may indeed be linked to the development of DME across the studies.

The results of our study indicate that individuals with hypertension had a 43% higher likelihood of experiencing DME compared to those without hypertension, Hypertension contributes to endothelial dysfunction and microvascular damage, exacerbating diabetic retinopathy and potentially DME. Effective blood pressure management is crucial in preventing and managing DME, emphasizing comprehensive cardiovascular risk management in diabetic care.

These findings align with a prior study on the relationship between blood pressure levels and diabetic retinopathy in patients with diabetes mellitus, which also found a significant association between hypertension and DME (14). This consistency suggests that hypertension is consistently linked to an increased risk of DME across multiple studies. And the finding from this study is different from study done in china where hypertension was not associated with macula edema and the different was explained by different study population.

Duration of diabetes mellitus (DM) of 5years or more was associated with a 47% more higher prevalence of DME in our study. Prolonged exposure to hyperglycemia leads to biochemical and structural changes in the retina, promoting inflammation, oxidative stress, and vascular permeability, all contributing to DME. Early diagnosis and aggressive management of diabetes are crucial in mitigating the cumulative impact of prolonged hyperglycemia on diabetic complications. Abnormal body mass index (BMI) was associated with a 46% higher prevalence of DME in our study. Obesity is a recognized risk factor for insulin resistance and systemic inflammation, which exacerbate diabetic microvascular complications like DME. Strategies for weight management and lifestyle modifications are integral to diabetic care, improving metabolic control and reducing the risk of DME.

One limitation of our study is its cross-sectional design, which restricts the ability to establish causality between identified risk factors and the development of diabetic macular edema (DME). Additionally, the reliance on self-reported data for certain variables such as medication adherence and lifestyle factors may introduce reporting bias. The study population, being drawn from a single geographic region, may not fully represent the diversity of type 2 diabetes patients globally, potentially limiting the generalizability of our findings.

## Conclusion

Our study highlights a significant burden of diabetic macular edema among type 2 diabetes patients, with an incidence rate of 33.9%. Key contributing factors identified include hypertension, serum cholesterol, low glomerular filtration rate abnormal body mass index, and duration of more than 5 years. These findings align with existing literature, reinforcing the importance of comprehensive diabetes management to moderate the risk of DME. The high prevalence of various types of macular edema, particularly diffuse and cystoid macular edema, underscores the need for tailored screening and intervention strategies. By addressing both metabolic and vascular risk factors, it is possible to reduce the burden of DME and improve visual outcomes for patients with type 2 diabetes.

## Data Availability

All data produced in the present work are contained in the manuscript

